# When can we safely return to normal? A novel method for identifying safe levels of NPIs in the context of COVID-19 vaccinations

**DOI:** 10.1101/2021.04.20.21255350

**Authors:** Gianluca Bianchin, Emiliano Dall’Anese, Jorge I. Poveda, Andrea G. Buchwald

## Abstract

Over the course of the COVID-19 pandemic, governing bodies and individuals have relied on a variety of non-pharmaceutical interventions (NPIs) to control the transmission of SARS-CoV-2, which posed an acute threat to individuals’ well-being and consistently impacted economic activities in many countries worldwide. NPIs have been implemented at varying levels of severity and in response to widely-divergent perspectives of risk tolerance. Now, concurrently with the introduction of multiple SARS-CoV-2 vaccines, the world looks optimistically to a “return to normality”. In this work, we propose a multi-disciplinary approach, combining transmission modeling with control and optimization theory, to examine how risk tolerance and vaccination rates will impact the safe return to normal behavior over the next few months. To this end, we consider a version of the Susceptible-Exposed-Infected-Recovered transmission model that accounts for hospitalizations, vaccinations, and loss of immunity. We then propose a novel control approach to calibrate the necessary level of NPIs at various geographical levels to guarantee that the number of hospitalizations does not exceed a given risk tolerance (i.e., a maximum allowable threshold). Our model and control objectives are calibrated and tailored for the state of Colorado, USA. Our results suggest that: (i) increasing risk tolerance can decrease the number of days required to discontinue all NPIs; (ii) increasing risk tolerance inherently increases COVID-19 deaths even in the context of vaccination; (iii) if the vaccination uptake in the population is 70% or less, then return to normal behavior within the next year may newly stress the healthcare system. Furthermore, by using a multi-region model accounting for travel, our simulations predict that: (iv) relaxation should take into account regional heterogeneity in transmission and travel; and (v) premature relaxation of NPIs, even if restricted only to low-density regions, will lead to exceeding hospitalization limits even when highly-populated regions implement full-closures. Although the simulations are performed for the state of Colorado, the proposed model of transmission and control methods are applicable to any area worldwide and can be utilized at any geographical granularity.

## 1 Introduction

The primary strategy for mitigating the spread of SARS-CoV-2 in the United States, to date, has relied on the use of non-pharmaceutical interventions (NPIs), including, at various levels of severity, city or state-wide lockdowns, travel restrictions, contact tracing, mask wearing, and social distancing. As an example, beginning March 2020, in the state of Colorado, United States, a variety of NPIs have been implemented, including stay-at-home orders, restrictions on indoor dining and domestic gatherings, business capacity limits, along with a state-wide mask mandate [1, 2]. Nation-wide lock-downs with stringent stay-at-home orders have been implemented in different countries such as Italy, United Kingdom, Israel, China, and Colombia, just to mention a few. These restrictions have had wide-ranging social consequences, including severe impacts on countries’ economies [3], and affected the well-being of families and children due to confinement stress and social disruptions [4]. At present, after most countries have recently initiated mass vaccination campaigns [5], individuals and policy-makers alike are planning a “return to normality”, where all NPIs can be gradually lifted and social behaviors can resume under circumstances that, theoretically, will not newly threaten the capacity of health-care systems.

On December 14, 2020 a mass vaccination campaign was initiated in the United States and, as of March 10, 2021, there are three vaccines officially approved by the U.S. Food and Drug Administration. Although current data on vaccine effectiveness is encouraging, ranging from ∼ 70% to ∼ 95% [6, 7, 8, 9, 10], vaccine hesitancy is becoming widespread [11, 12]. Vaccinations effectively help to contain the spread of an epidemic by quickly developing immunity in vaccinated individuals, without causing severe illness. As the majority of a population becomes immune to SARS-CoV-2 thanks to vaccinations, the population should rapidly reach a condition in which each infectious individuals can transmit the infection, on the average, to a number of individuals that is strictly smaller than one, thus preventing new outbreaks. As the spread of the virus is slowed by aggressive vaccination campaigns, NPIs can be gradually lifted and social interactions can partially resume in the interest of re-establishing economic and social activities.

Removal of NPIs and resumption of normal behavior should ideally occur only when a “high-enough” fraction of the population is immune in order to prevent additional waves of infection, which may newly overwhelm the healthcare system. However, predicting what is the minimum level of immunity that is needed to remove all NPIs remains an open research question, especially in the presence of regional discrepancies in risk tolerance and vaccination rates. For example, although in the state of Colorado many NPIs have been introduced in a top-down fashion, at the county-level there are several local variation in restrictions, dependent on local risk tolerance and transmission metrics. This has led to a patchwork of interventions that reflects the widespread variation in control measures nationally and globally. Removal of NPIs will likely continue to occur in a patchwork fashion, dependent on local risk tolerance and vaccination rates.

Motivated by the previous background, in this paper we aim to answer the following questions: given the ongoing vaccination campaigns and a range of hospitalization risk tolerance, when can all NPIs be repealed such that life can safely return to normal? How do travel patterns between different regions impact the return to normal? And, lastly, how is this return to normal impacted by variations in vaccination rates and risk tolerance in a heterogeneous but connected region? To answer these questions, we use a novel mathematical model that brings together transmission modeling and advances in optimization theory and feedback control. The proposed framework is engineered to select optimal allowable levels of transmission-relevant contacts across geographical regions based on current vaccination rates, estimated infection rates, and a maximum allowable hospitalization limit (representing levels of risk-tolerance). The latter aspect is of upmost importance to maintain stability of the health-care system and ensure that future waves of infections do not threaten public health infrastructures. In particular, our approach entails the following efforts:

i. We consider an extension of the Susceptible-Exposed-Infected-Recovered (SEIR) transmission model that accounts for hospitalizations and vaccinations. We consider two variations of this model: a single-region model, which describes the behavior of the entire population at the aggregate-level neglecting regional diversities, and a more-comprehensive network model that takes into account structured coupling between regions.
ii. We leverage recent advances in feedback-based online optimization of dynamical systems [13, 14, 15, 16, 17] to synthesize a control algorithm that selects the level of transmission-relevant contact (hereafter referred to as contact level) needed under varying levels of tolerance in hospitalizations. Here, the objective of the control policy is to minimize the severity of the NPIs and behavioral modifications (i.e., to minimize the consequences on economic, as well as social, and individual-level health) while ensuring that a pre-specified hospitalization limit is not exceeded. From a mathematical standpoint, the proposed controller builds upon the formulation of an optimization problem that captures the constraints on the number of hospitalizations or peak infections, as well as severity of NPIs.

We use the state of Colorado, USA, as a case-study to test the performance of the controller and to show how risk tolerance and vaccination rates impact the strategy and timing of safe return to normal. In our experiments, we consider a strict constraint on the maximum number of allowable hospitalizations, and we show how this parameter affects the timing of safe return to normal. Although the results are provided for the state of Colorado, the proposed methods are widely applicable to investigate the relationship between risk tolerance, vaccination rates, and NPIs. The methods can be applied at different geographical scales, based on how NPI decisions are split across state, regional, or county-wide governing bodies.

## 2 Results

### Controller Structure and Model Description

We derived a feedback controller to select an optimal level of allowable transmission-relevant contacts that balances between the economic impact of NPIs and the number of infected individuals at the endemic equilibrium, while simultaneously guaranteeing that the maximum number of daily hospitalized individuals is bounded by a pre-specified limit at all times (see Methods for a description of the control method). The technique is inspired by online projected gradient dynamics for constrained optimization problems. We apply the proposed controller to a compartmental model that incorporates hospitalizations, vaccinations, and loss of immunity. The model includes seven compartments: Susceptible, Exposed, Infected, Hospitalized, Recovered, Vaccinated, and Deceased. The adopted model is described in detail in Section 4 (see also Figure 7 for an illustration). The effects of NPIs are abstracted mathematically by introducing a control parameter *u* ∈ [0, 1] that models the level of permitted transmission-relevant contacts, where *u* = 0 corresponds to a full lockdown (zero contacts) and *u* = 1 corresponds to absence of any NPIs or behavioral changes (pre-pandemic contact levels). The control method incorporates a parameter *h*_lim_ that describes the maximum number of allowed hospitalized individuals.The proposed control framework automatically updates the intensity of NPIs in order to guarantee that the number of hospitalized individuals at every time does not exceed the limit *h*_lim_. To assess the economic costs of state-wide and regional-level NPIs, we use the convex loss function proposed in [18] that was adopted to model the cost of lockdown in the state of New York, USA.

As further described in Methods, the model was fitted to data from the state of Colorado [19], USA. Precisely, the model parameters were chosen to fit hospitalization data for the period from January 1st, 2021, to February 28th, 2021. Hospitalizations declined steadily during this time.

The control technique is then extended to capture regional diversity by using a networked version of the model. The network model is fitted to a regional resolution where the state is partitioned according to Local Public Health Agency (LPHA) regions [20]. Similarly to the single-region scenario, each region features a control variable *u*_*i*_ that describes the level of permitted social transmission-relevant contacts within region *i*. Mobility data from cell-phone usage obtained from SafeGraph (https://docs.safegraph.com/docs/social-distancing-metrics) is used to estimate interaction matrices describing inter-regional couplings (see Methods).

### Assessing Risk Tolerance by Limiting the Number of Allowed Hospitalizations

We first assess the efficacy of the proposed control method by focusing on the problem of selecting the least-restrictive level of NPIs that guarantees that the number of hospitalized individuals does not exceed the threshold *h*_lim_. Namely, we first focus on the scenario where no penalty is assigned to a high number of infected individuals at the endemic equilibrium.

Simulation results in Figure 1 illustrate the trajectories of transmission-relevant contacts, hospitalizations, and infections for the single-region model parametrized for the entire state of Colorado. In these simulations, in order to showcase the benefits of the control method, we assume that vaccinations are unavailable. Absence of vaccinations is chosen as a base-case to evaluate the progression of the epidemic and the effectiveness of the NPI controller. We consider three scenarios, where the state-wide hospitalization limit takes the values 300 individuals/day, 500 individuals/day, and 1200 individuals/day, respectively. In the first two cases, the method predicts that the least-restrictive level of transmission-relevant contacts that allows the model to abide the hospitalization limit is *u* = 0.269 and *u* = 0.312, respectively. This is comparable to a 73% and 69% decrease in total transmission-relevant contacts comparable to pre-pandemic behavior, respectively. With a higher risk tolerance of 1200 hospitalized individuals/day, the controller allows a gradual increase in contact levels from 30% on 4/20/21 to 67% on 2/14/22. Notice that, in this case, the allowable levels of transmission-relevant contacts can be gradually increased because the population quickly-transfers to the Recovered compartment due to naturally-acquired immunity. Precisely, in this case 50% of the population is in the Recovered compartment by 9/27/21.

**Figure 1:**
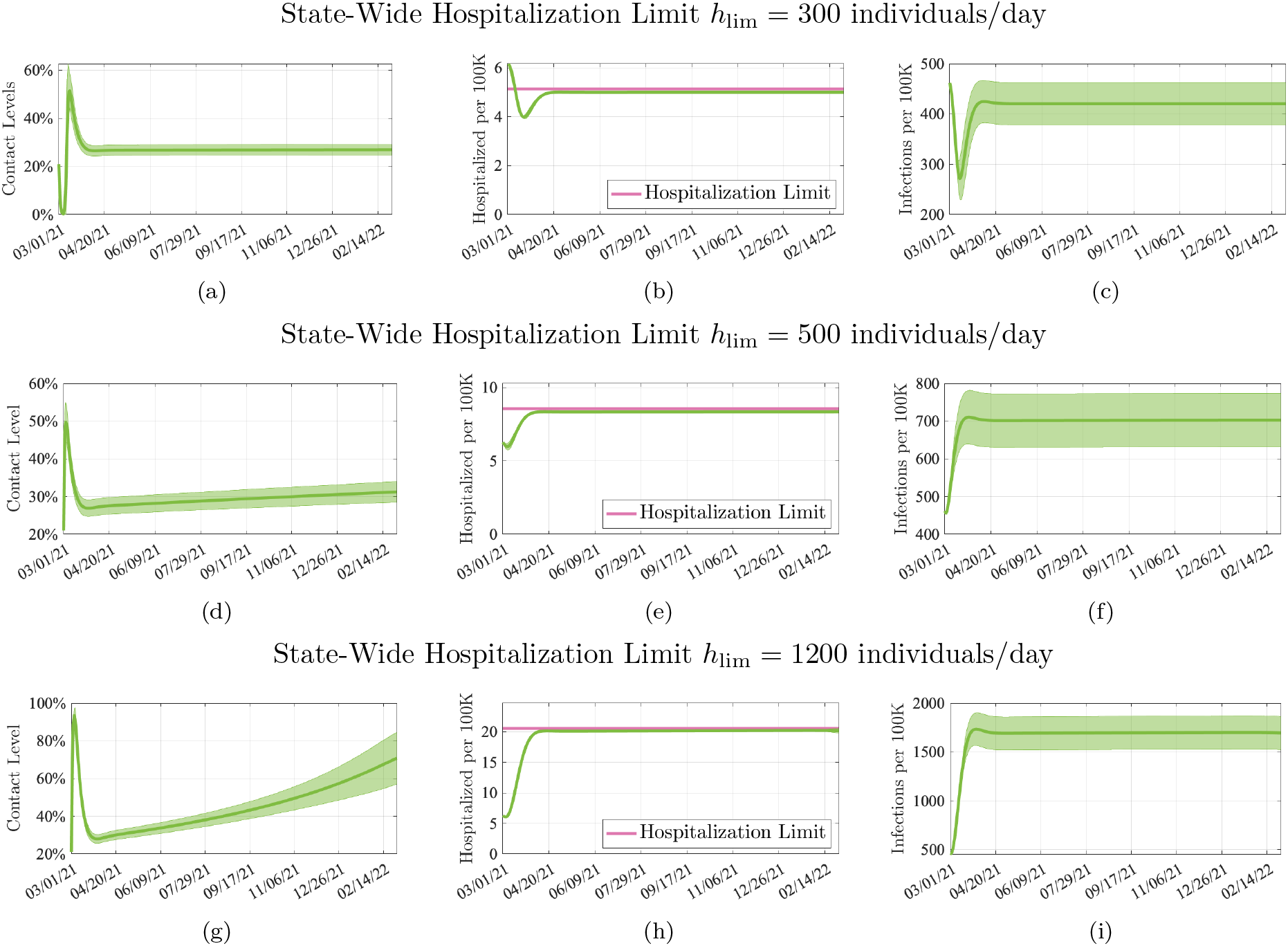
Model behavior when minimizing the severity of NPIs while bounding the maximum number of allowed hospitalizations. Each row of panels illustrates a different choice of hospitalization limit. All simulations are conducted by assuming absence of vaccinations (to provide a base case), and focus on a single-region model that is fitted using data from the state of Colorado, USA (see Section 4). Results are averaged over 10,000 simulations with parameters sampled using a Latin Hypercube technique around their nominal values. The solid line shows the mean of the trajectory, and the shaded areas show 3 − σ confidence intervals. (a)-(c) Hospitalization limit = 300 individuals/day. (d)-(f) Hospitalization limit = 500 individuals/day. (g)-(i) Hospitalization limit = 1200 individuals/day.

As shown in Figure 1(b), when the number of hospitalized individuals is larger that the specified limit, the controller quickly steers the number of hospitalizations down to acceptable levels by reducing the allowable levels of transmission-relevant contacts *u*. After the number of hospitalized individuals is below the specified limit, the controller guarantees that the constraint is satisfied at all future times, as shown in Figure 1(e)-(h). Figure 1(c)-(f)-(i) suggest that maintaining the number of hospitalizations near a specified threshold at all times implies that the number of infections also remains approximately bounded. Overall, Figure 1 demonstrates that the proposed control method is effective in maintaining the number of hospitalizations below (and near) the pre-specified threshold.

Figure 2 illustrates the impact of different vaccination rates in the required level of NPIs. Similarly to the simulation in Figure 1, the controller is calibrated to minimize the severity of the NPIs, while guaranteeing that the number of hospitalizations is below the pre-specified limit. Our models suggest that, when the vaccination rate (for the entire state of Colorado) is 15, 000 vax/day, the allowable contact levels can gradually return to normality, i.e., the control variable (Figure 1(a) and (e)) can be gradually increased from *u* = 0.286 on 4/1/21 to *u* = 0.975 in 320 days (i.e., on 2/15/22). In contrast, when the vaccination rate is increased to 25, 000 vax/day, NPIs can be gradually lifted from *u* = 0.286 on 4/1/21 to *u* = 1 after 193 days (i.e., on 10/11/21). Notice that, in this simulation, we used an ideal vaccination uptake of 100% as shown in Figure 2(h). Although such vaccination uptake may be impractical, this ideal situation was utilized to provide a best-case analysis. These simulations should be compared with Figure 4 (discussed below) where the role of different vaccination uptakes is analyzed.

**Figure 2:**
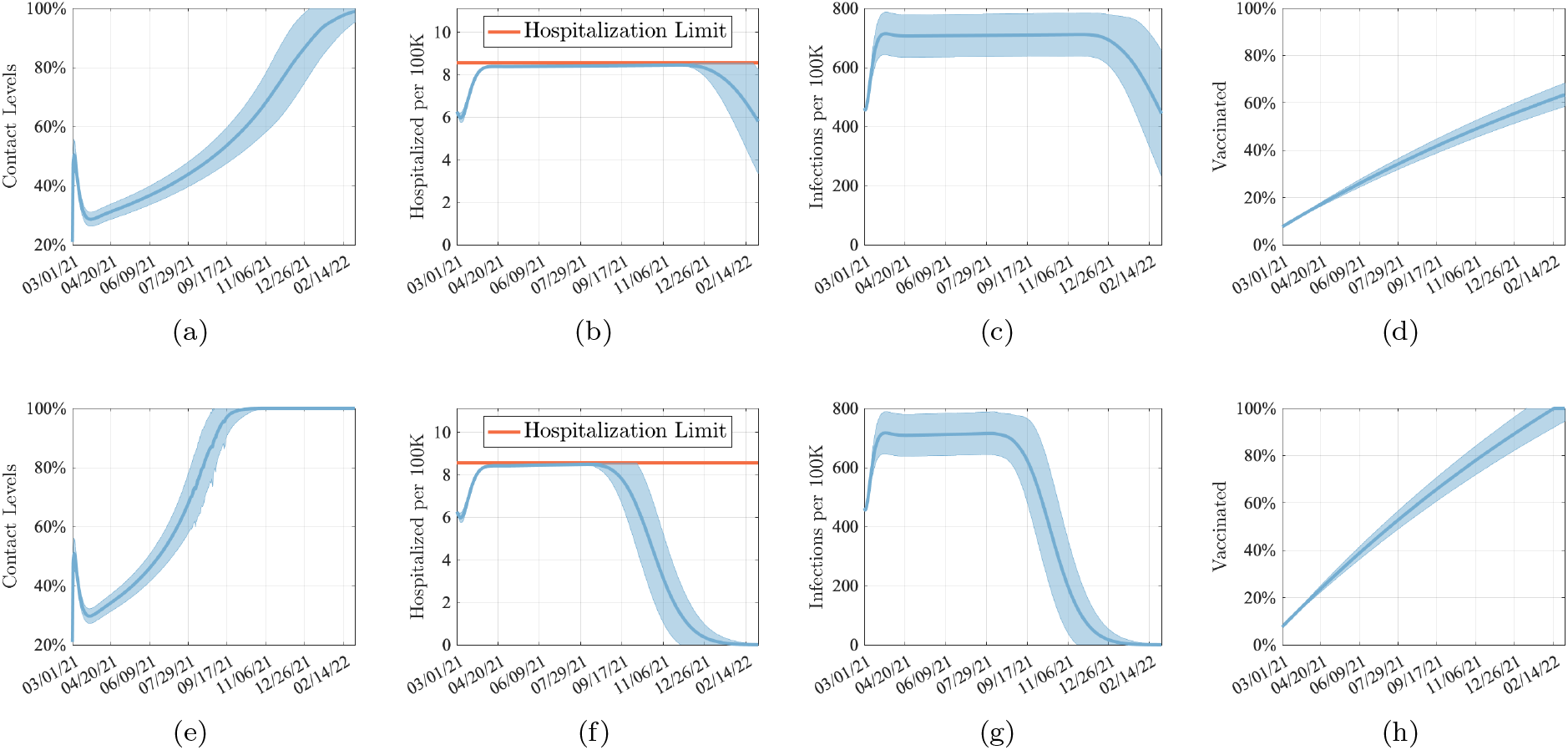
Model behavior when minimizing the severity of NPIs while bounding the number of allowed hospitalizations. Each row of panels considers a different vaccination rate. All simulations are conducted by considering a hospitalization limit of h_lim_ = 500 individuals/day, and focus on a single-region model that is fitted using data from the state of Colorado, USA (see Section 4). Results are averaged over 10,000 simulations with parameters sampled using a Latin Hypercube technique around their nominal values. Continuous line shows mean of the trajectory and shaded area show 3 − σ confidence intervals. (a)-(d) Vaccination rate = 15, 000 vax/day. (e)-(h) Vaccination rate = 25, 000 vax/day.

Figure 3 illustrates the implications of considering combined control objectives. Figure 3(a)-(d) illustrates the model behavior when the controller is calibrated to simultaneously minimize the severity of NPIs and minimize the number of infections at the endemic equilibrium (i.e., the number of infected individuals at time *t* = ∞). Such strategy seeks to find a balance between the economic impact of NPIs and the social risk associated with the number of infections in the long run. Notice that these simulations have been performed over a time horizon of 4 years in order to illustrate the properties of the endemic equilibrium. In contrast, Figure 3(e)-(h) illustrates the epidemic evolution when the controller only minimizes the severity of NPIs. For this simulation, we considered a vaccination uptake of 75% and we set the limit of allowed hospitalizations to 500 individuals/day. By comparing Figure 3 (b) and (f), our models suggest that in both simulation scenarios the controller guarantees that the number of hospitalizations does not exceed the hospitalization limit. By comparing Figure 3 (c) and (g), our models suggest that when the controller balances between the severity of NPIs and simultaneously seeks to reduce the number of infections at the endemic equilibrium, the number of infections on 2/28/25 is 482 individuals per 100K, which represents a 17% reduction with respect to number of infections when the controller only optimizes the severity of NPIs. As illustrated by Figure 3(e), when the controller’s objective is to solely minimize the severity of NPIs, all restrictions on allowable levels of transmission-relevant contacts are entirely lifted by 07/14/22. On the other hand, when the controller seeks to balance the severity of NPIs with the number of infections, the allowable levels of transmission-relevant contacts assumes values no larger than 80% in order limit the number of infections.

**Figure 3:**
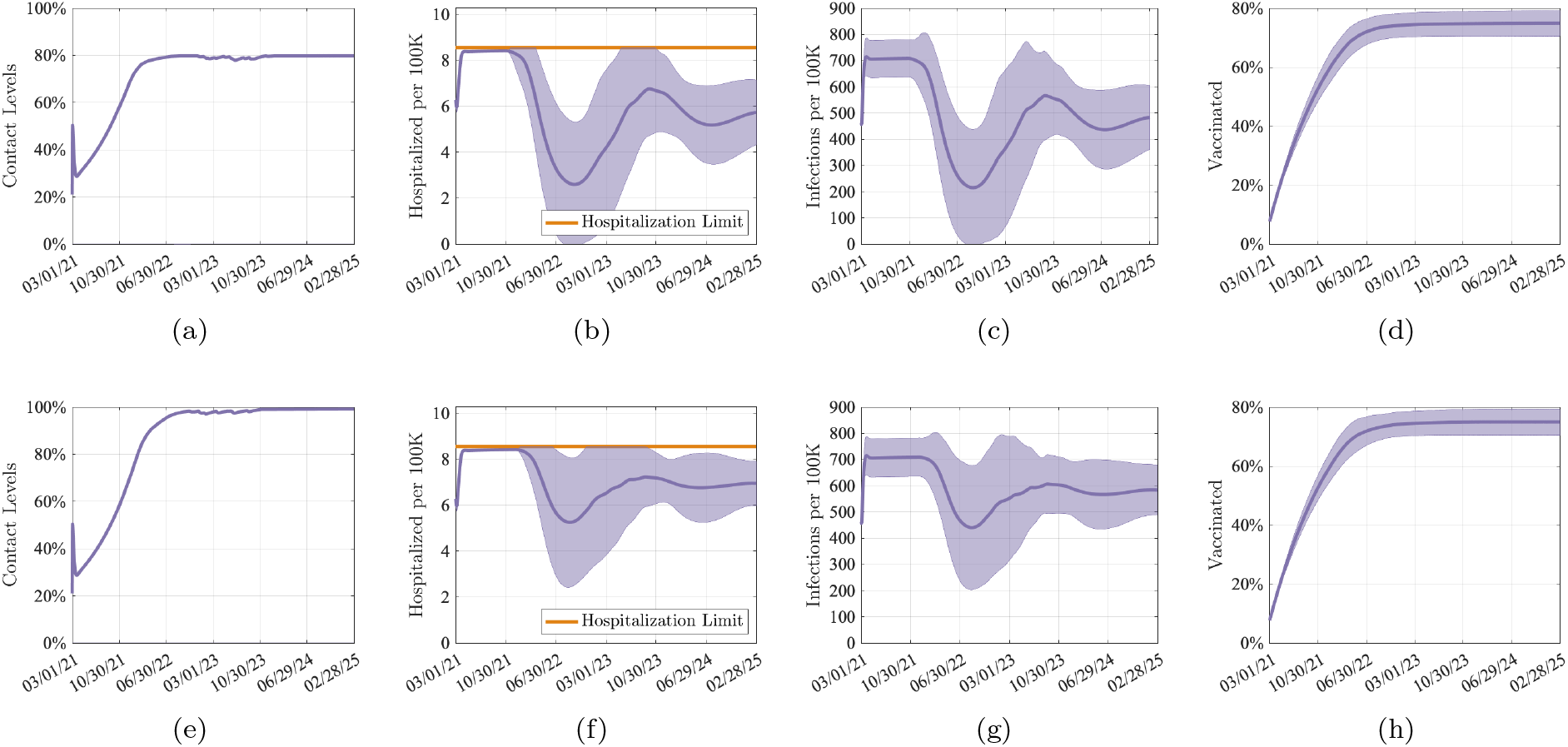
Model behavior under the assumption of 75% vaccine uptake and 500 hospitalized individuals/day when: (a)–(d) the controller optimizes both the number of infections at the endemic equilibrium and the severity of NPIs, and (e)–(h) the controller minimizes only the severity of NPIs. Results are averaged over 10, 000 simulations with parameters sampled using a Latin Hypercube technique around their nominal values. The continuous line shows the mean of the trajectory, and the shaded area shows 3 − σ confidence intervals.

### Return to Normality

The availability of a feedback-control policy that can select the least-restrictive level of NPIs to satisfy a hospitalization limit allows us to determine a lower bound to the number of days required for a return to normality; namely, the number of days before all NPIs can be removed (i.e. *u* = 1). Figure 4 shows the number of days (beginning 03/01/21) that are necessary before a full return-to-normality can be implemented, while ensuring that the given hospitalization limit is not exceeded. Figure 4(a) shows the number of days to normality in relationship to different choices of hospitalization limits. The method predicts that, with a vaccination rate of 21, 000 vaccines/day, the number of days before all NPIs can be lifted reduces by a factor of two (i.e. from 300 days to 164 days) as the number of allowed hospitalizations is increased from 300 individuals/day to 1, 200 individuals/day. We note that, although the number of days to normality is a decreasing function of the hospitalization limit, increasing the hospitalization limits results in a higher number of deaths, as shown in Figure 4(c). This behavior originates because, as the number of allowed hospitalizations is increased, more individuals achieve natural immunity (i.e. through symptomatic infection). Figure 4(b) illustrates the number of days before all NPIs can be lifted in relationship to different levels of vaccine uptake. When the hospitalization limit is maintained below 400 individuals/day, the method predicts that more than 700 days are necessary to lift all NPIs, when the vaccination uptake is no larger than 60%. The model also predicts that a vaccination uptake of 70% is necessary to lift all NPIs in less than 700 days when the hospitalization limit is set to no more than 300 individuals/day. Figure 4(c) illustrates the cumulative number of deaths from 03/01/21 for different levels of allowed hospitalizations and vaccination rates. Figure 4(c) shows that the number of deaths grows linearly with the number of allowed hospitalizations (precisely, the number of deaths grows with slope 1.9 as the number of allowed hospitalizations increases); in contrast, the simulations suggest that the number of deaths is independent of the rate of vaccinations. This behavior can be interpreted by noting that the number of deaths is proportional to the number of infections and of hospitalized individuals, which are maintained constant over time by the control method.

**Figure 4:**
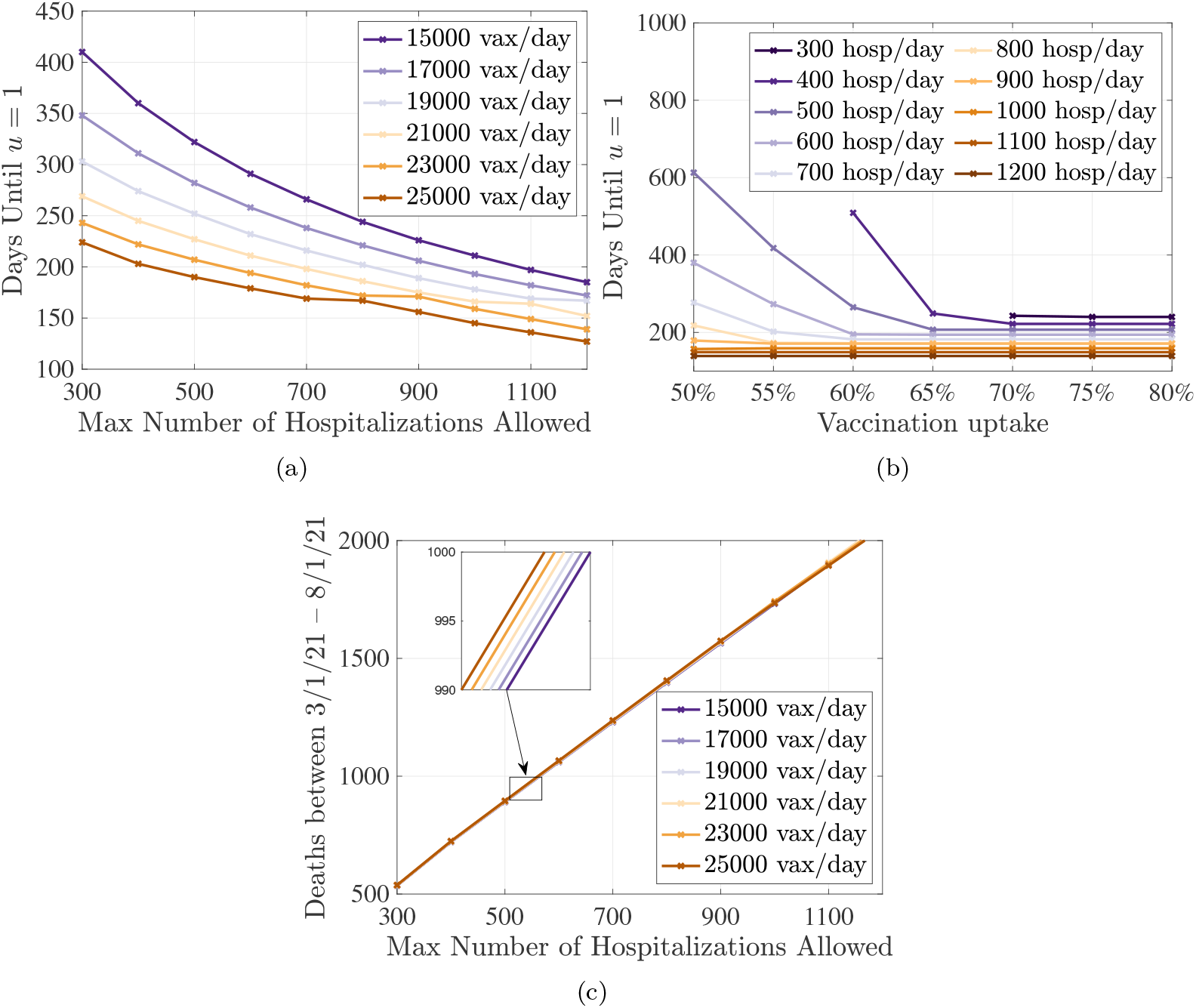
Number of days to normality (i.e. all NPIs can be lifted) beginning 3/1/21 as a function of hospitals’ risk tolerance, vaccination rate, and vaccination uptake. These results are obtained in the case where the objective is to minimize the severity of NPIs, subject to a limit on the hospitalizations. (a) Number of days to normality as a function of the limit of hospitalizations, for different rates of daily vaccinations. (b) Number of days to normality as a function of the vaccination uptake, for different hospitalization limits. Vaccination rate is y = 23, 000 vax/day. Data points for 300 hosp/day and 400 hosp/day and vaccination uptake lower than 60% and 70%, respectively are not displayed because u did not reach 1 for the duration of the simulation. (c) Estimation of the number of deaths between March 1, 2021, and August 1, 2021, as a function of the limit in the hospitalizations and different rates of daily vaccinations.

### Regional Effects

A crucial open question concerns how to support local authorities in selecting optimal, local-level NPIs in order to prevent state-wide lockdown. To this aim, we adapted the model to account for regional variations in transmission using a network model fitted to hospitalization for the period from January 1st, 2021, to February 28th, 2021 for 11 LPHA regions in Colorado [21]. See Figure 11 for an illustration of the LPHA-based regional partitioning for the state of Colorado and Table 1) for a summary of the regions. We used the proposed feedback-based controller to select optimal regional NPIs in order to guarantee that the number of hospitalized individuals in each LPHA region satisfies pre-specified, region-dependent, hospitalization limits. Regional hospitalization limits have been obtained by normalizing the state-wide limit of 500 individuals/day by the size of the populations in each LPHA region and rounded to the nearest integer towards +∞. Figure 5 shows the number of hospitalized individuals in the time-interval 03/01/21 – 03/01/22 as well as the optimal level of control needed to satisfy the regional hospitalization limits. By comparing the simulation outcomes across the various regions, our model and control methods suggest that there is a high-discrepancy in the least-restrictive level of NPIs required across the regions. Indeed, values range from *u*_*i*_ ≈ 1 (equivalent to a full removal of all NPIs) in the Central Mountains region on 07/01/21, to *u*_3_ ≈ 0 (equivalent to a complete lockdown) in the East Central region on the same date. By observing that *u*_3_ ≈ 0 is a practically unrealistic situation, this simulation outcome suggests that a proportional-allocation of critical care units inherently originates unfairness in the required intensity of NPIs across the regions, and that, instead, the allocation of critical care units should also account for the network connectivity of each region.

**Table 1:**
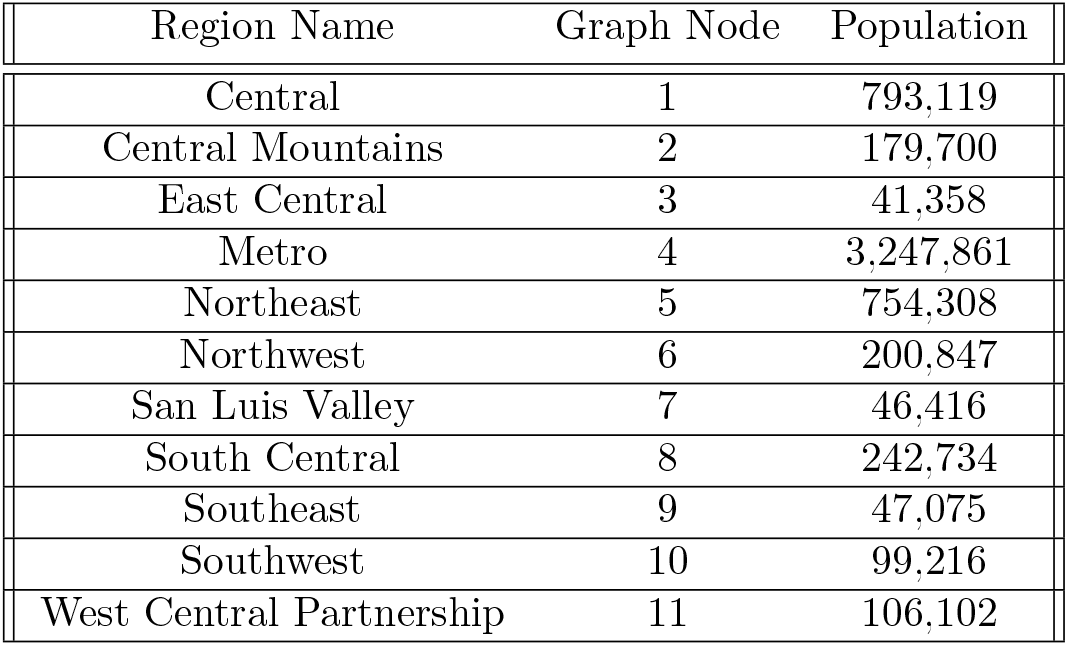
Properties of LPHA regions.

| Region Name | Graph Node | Population |
| --- | --- | --- |
| Central | 1 | 793,119 |
| Central Mountains | 2 | 179,700 |
| East Central | 3 | 41,358 |
| Metro | 4 | 3,247,861 |
| Northeast | 5 | 754,308 |
| Northwest | 6 | 200,847 |
| San Luis Valley | 7 | 46,416 |
| South Central | 8 | 242,734 |
| Southeast | 9 | 47,075 |
| Southwest | 10 | 99,216 |
| West Central Partnership | 11 | 106,102 |

**Figure 5:**
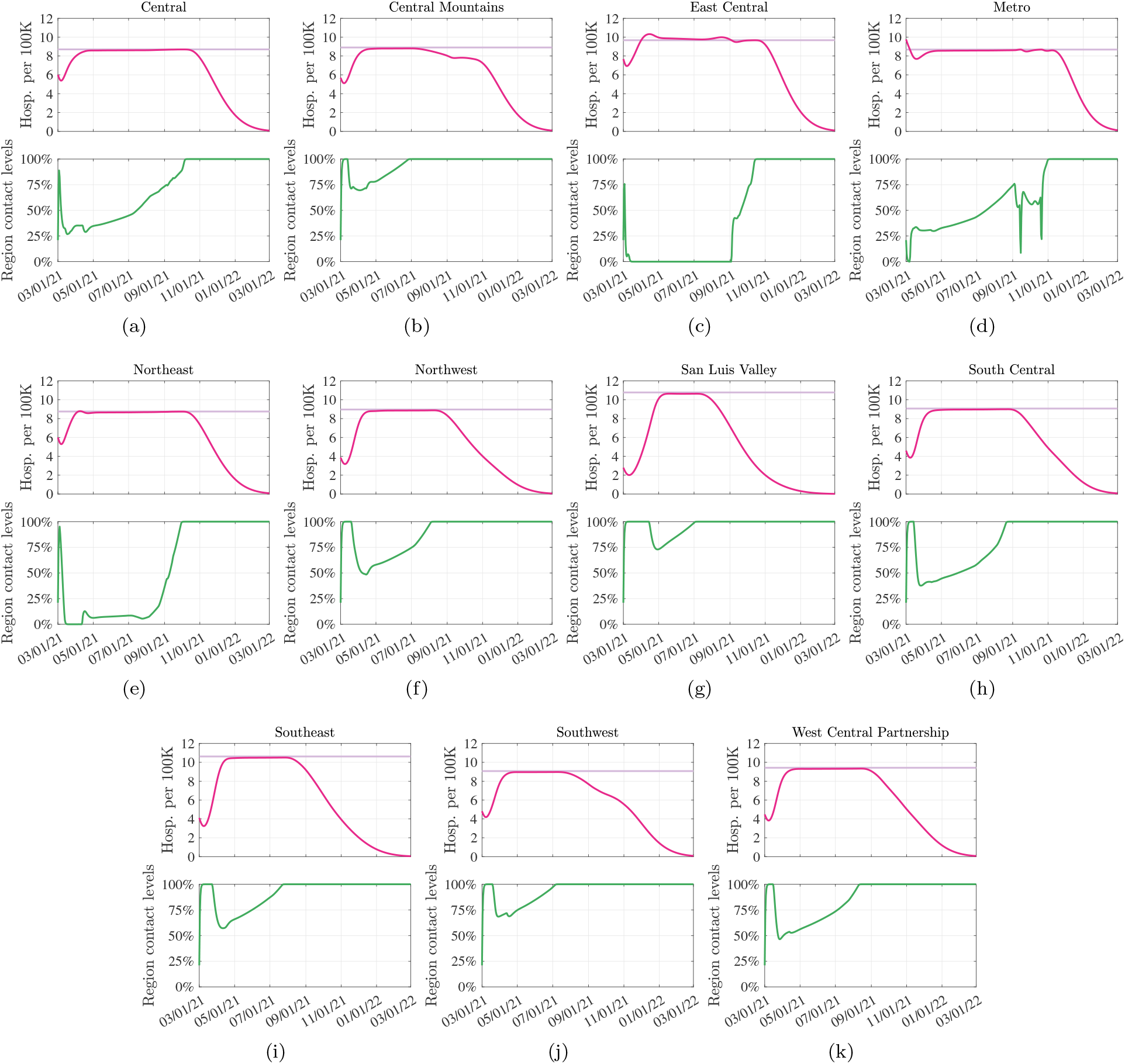
Hospitalizations and controller level over time when a group of regional controllers are used to guarantee that pre-specified region-dependent hospitalization limits are not violated. Each of the 11 panels shows the evolution in a different LPHA region (see Figure 11 for an illustration of the connectivity graph). Simulation conducted with vaccination rate is y = 20, 000 vax/day. The scale of the vertical axis (top plots) is hospitalization/day per 100K inhabitants. Dark purple lines show the evolution of hospitalizations, light purple lines illustrate the pre-specified hospitalization limit. Solid green lines illustrate the required level of NPIs u_i_.

Figure 6 investigates a scenario where regions with a population of 150, 000 people or less (i.e., East Central, San Luis Valley, Southeast, Southwest, and the West Central Partnership regions) drop most NPIs as of 05/01/21, to a level of *u* = 0.8. On this date, the average fraction of fully vaccinated individuals across the state is 21.29%. As illustrated by the simulation, such policy will result in a violation of the hospitalization limit in 7 out of 11 regions (i.e. Central Mountains, East Central, Metro, Northeast, Southeast, Southwest, and West Central Partnership). Interestingly, our results suggest that even if the highly-populated regions Metro, Central Mountains, and Northeast implement a complete lockdown (i.e., *u*_*i*_ ≈ 0) immediately after 05/01/21, their hospitalization limits will be violated beginning 05/13/2021. Not surprisingly, the regions that decrease NPIs too soon are also the ones that are affected by the highest number of hospitalizations, with peaks around June 25, 2021.

**Figure 6:**
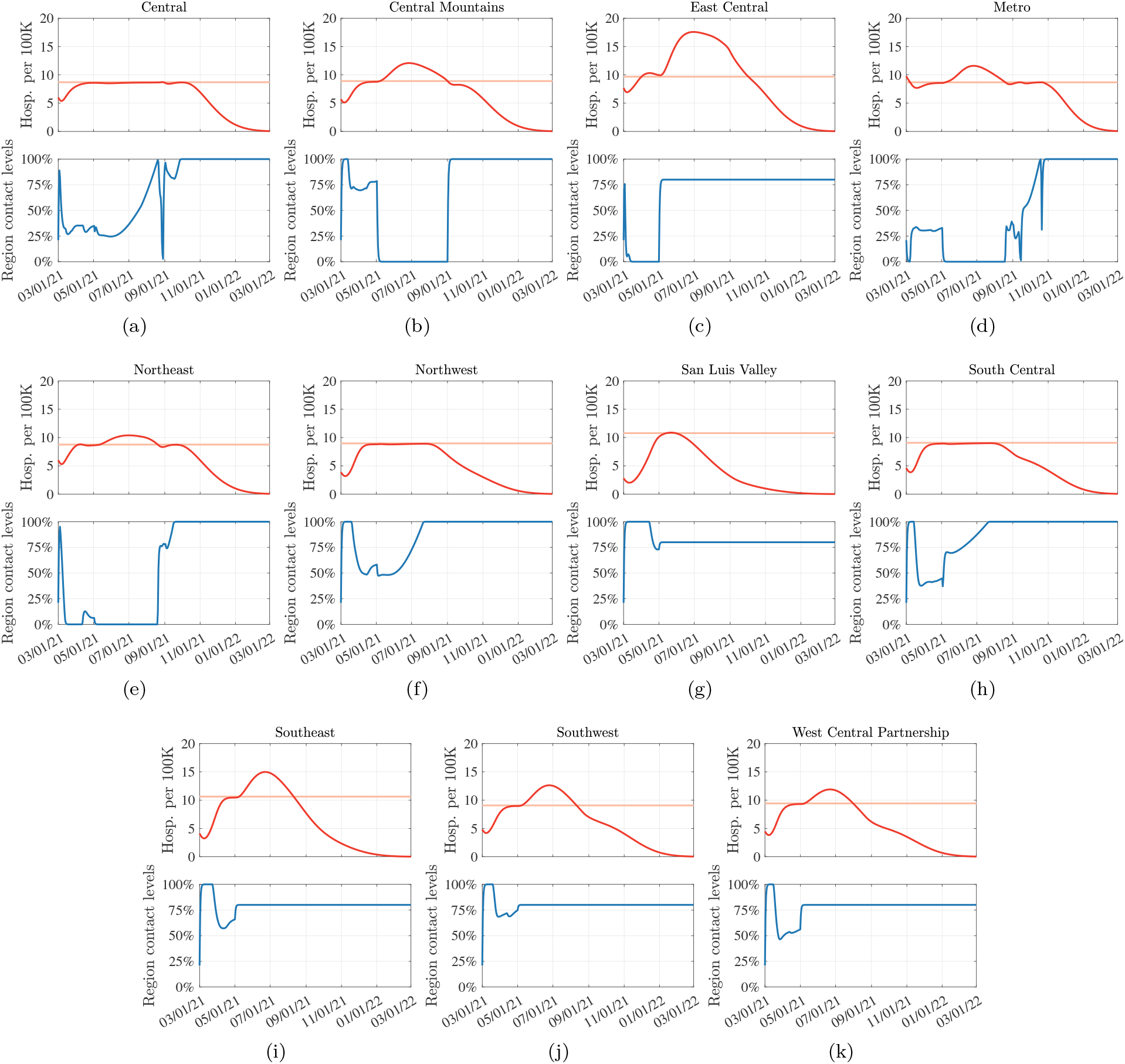
Hospitalizations and controller level over time when regions with a population of 150, 000 people or less (i.e., East Central, San Luis Valley, Southeast, Southwest, West Central Partnership) drop all NPIs on 05/01/21. Simulation conducted with vaccination rate is y = 20, 000 vax/day. Each of the 11 panels shows the evolution in a different LPHA region (see Figure 11 for an illustration of the connectivity graph). Top plots show hospitalization data, bottom plots illustrates the level of NPIs that guarantees that regional hospitalization limits are not violated. The scale of the vertical axis (top plots) is hospitalization/day per 100K inhabitants. Dark red lines show the evolution of hospitalizations, orange lines illustrate the pre-specified hospitalization limit.

We conclude by noting that, although the simulations in Figure 6 are performed for the state of Colorado as an illustration, the proposed model and controller are applicable to different geographical areas and can be implemented at a different geographical granularity. For example, one may model the evolution of the epidemic in the United States of America, where each region in the model corresponds to a state.

## 3 Discussion

Despite optimism over widespread vaccination, a safe return to resuming pre-COVID contact behaviors, without any NPIs, may still be a long way away, dependent on the number of SARS-CoV-2 infections and consequent severe COVID-19 cases we are willing to tolerate. Using a novel method combining a deterministic compartment model with a feedback-based optimization method, we examine the conditions under which all NPIs can be removed and individuals can resume pre-pandemic contact behaviors. We successfully show that the proposed controller can be used to indicate necessary increases or decreases in NPIs based on the level of community risk tolerance or how severe the hospitalization burden can be before governments will act to impose restrictions or individuals choose to change their behaviors. While increasing vaccination rates will lead to a decreased “time to normal,” under low levels of risk tolerance safe return to normal may not occur until early 2022. Allowing for increased burden of hospitalization decreases the time to a safe return to normal behavior, but with serious consequences in the form of increased morbidity and mortality, even under scenarios with high vaccination rates. Regional heterogeneity may complicate this picture and low-population regions with relatively low contact rates may begin to return to relative normality far more quickly. However, due to high connectivity, removing NPIs too quickly in regions of low population density can still lead to dire consequences in nearby high-density regions.

For the purpose of this study we utilize a single control variable to encompass all policy measures and behavioral changes in a region that decrease transmission of SARS-CoV-2, excepting vaccination. This variable is assumed to capture the impact of policy measures; these includes mask mandates, school closures, business capacity limits, as well as personal decisions such as hand-washing, mask wearing, and moving socialization outside that otherwise would have occurred inside. We do not attempt to model individual policy measures or behaviors due to the fact that implementation of multiple NPIs and behavioral changes have occurred simultaneously and the individual effects are difficult to disentangle [22]. While NPIs implemented by policy-makers are important, a large portion of transmission reducing behaviors are a result of individual-level risk assessment and behaviors, in response to perceived community transmission [23]. Thus, even as policy makers begin to relax NPIs at the state and regional levels, individuals will continue to make decisions based on their own perceptions of risk, which are directly impacted by hospitalizations and infections in the community. As a result of these effects, we recognize that an actual implementation of various NPIs may have a high variance. For example, even if NPIs are lifted, as prevalence remains high, it might be unlikely that some individuals will return to pre-pandemic behavior. Likewise, even at the height of restrictions, when stay at home orders were in place, it may not be possible to control transmission entirely given the necessity of ongoing essential work, grocery shopping, etc (therefore, *u* can approach 0, but cannot be set to *u* = 0 in practice), or because of possible violations of restrictions.

### Differences relative to prior works

In this work, we take a novel approach to modeling the control of epidemics, based on the use of online feedback-based optimization methods for dynamical systems (see the previous works of the authors [14, 16, 17]). The proposed method takes as an input the number of infected and hospitalized individuals, along with a function that estimates the number of peak hospitalization based on the current model of the pandemic. This information is used by the proposed controller to adjust in real time the level of NPIs to meet economic and risk-related objectives; the objectives are encapsulated into an optimization problem, and the controller is designed to control the progression of the epidemic to meet those objectives. We propose controllers based on a single region, and also based on a network model of the epidemic. In the latter, each region implements a local controller for the NPIs. This work is broadly generalizable to other variations of epidemic models, including numerous variations on the well-studied SEIR model (properly modified to account for hospitalizations or other health-related metrics of interest), or the recently-proposed model SIDARTHE [24].

Although optimal control of epidemics under limited control resources is a classical problem [25, 26, 27, 28, 29, 30, 31], this well-investigated topic has recently gained renewed attention in view of a modern light in the context of control of the COVID–19 pandemic. As representative examples, an optimization-based approach is utilized in [18] to minimize the cost of a lockdown, while guaranteeing that the average number of infections decreases with a given rate; a model predictive control method is presented in [32]; optimal control problems are explored in, e.g., [33, 34, 35], and optimization approaches are explored in [36] to reduce economic losses with targeted closures. Timing of the social distancing was investigated in, e.g., [37]; NPI tuning starting from a dynamic model for social distancing based on the SIR epidemics model was proposed in [38]. Our work takes a different approach, based on the use of online feedback-based optimization methods for dynamical systems. Recent research efforts also proposed to control the epidemic evolution by selecting the least-restrictive level of NPIs that simultaneously maintains the number of infections below a given threshold and minimized the time required to achieve herd immunity [39]. Unfortunately, classes of strategies that rely on naturally-induced immunity originate a worse-case number of deaths. Here we extend these strategies accounting for vaccination and attempt to account for reasonable ranges of probable risk tolerance thresholds.

### Limitations

We acknowledge that our findings come with a number of important limitations. Our findings are dependent on numerous assumptions about baseline transmission rate, probability of hospitalization, and other parameter values estimated from previous modeling studies that may impact our results. For simplicity, we chose not to account for age or the differences between asymptomatic and symptomatic transmission which may have altered our findings. Despite accounting for regional heterogeneity in contact rates and baseline transmission, superspreader events and smaller non-homogenous spatial units play a large role at this stage in the pandemic. Vaccine distribution is occurring in a manner which reinforces pre-existing health disparities, due to issues of both access and hesitancy [40]. This creates pockets of high-risk unvaccinated populations, which are sufficient to sustain transmission, even with high vaccination rates overall. Our model cannot account for this type of clustering of behavior or risk, which are important in understanding the probability of achieving sufficiently low levels of SARS-CoV-2 transmission. Additionally, we do not account for the recent introduction and proliferation of numerous variant strains which have the potential to substantially alter transmission dynamics and vaccine efficacy [41]. Over time, current vaccines may be less effective at preventing infection due to new circulating variants, preventing attainment of herd immunity even with high rates of vaccination uptake. Similarly, the duration of immunity obtained from current vaccines against currently circulating variants is unknown. For the purpose of this study, we assumed an effectiveness of two years (which may be overly optimistic). If the duration of vaccine-derived immunity is short, given feasible vaccination rates, complete relaxation of NPIs may never be attainable [42]. When information and models on loss of immunity are available, the model utilized here can be modified accordingly. We also acknowledge that it may be difficult to directly translate a variable between 0 and 1 into precise actions such as mask mandates, school closures, and business capacity limits, and especially personal decisions; however, current studies are looking at regressing the reproduction number of COVID-19 against different NPIs [43], and this can naturally be connected to the variable *u* of the controller.

### Conclusions

Our findings are in agreement with previous modeling studies which have stressed the need to maintain current levels of NPIs and decreased contact for the near future, even in the context of current vaccination strategies [44, 45, 46]. Studies have additionally questioned whether herd immunity through vaccination is achievable at all, given the current vaccines available and the high prevalence of vaccine hesitancy [47]. Given these factors, the possibility has been raised that SARS-CoV-2 will become an endemic virus circulating regularly in the population [48] and our concept of a “return to normal” will have to be reframed. Currently (04/09/2021), in the state of Colorado, transmission has flattened with approximately 400 individuals currently hospitalized. Vaccination rates are high, however, whether or not we can reach 70% vaccine uptake is uncertain. Regardless of whether a true return to pre-pandemic behavior is achievable, we present herein a novel method for examining the necessary levels of NPIs under various levels of risk tolerance which can be used to determine safe levels of relaxation as well as estimate relative time-frames for policy and behavior changes.

## 4 Methods

In this section, we discuss the Susceptible-Exposed-Infected-Hospitalized-Recovered-Vaccinated-Susceptible (SEIHRVS) transmission model, and we describe the proposed NPI controller and discuss its implementation.

### 4.1 Single-Region Modeling and Control

We first consider a model of transmission defined for a given geographical region. The following sections will describe the SEIHRVS model, and will explain how the controller for NPIs is designed. After this, Section 4.2 elaborates on the network model.

#### 4.1.1 Scalar SEIHRVS Model

We model the progression of an epidemic in a population by using a variation of the Susceptible-Exposed-Infected-Recovered (SEIR) model that accounts for hospitalizations, vaccinations, and loss of immunity. In particular, we consider a transmission model with states: Susceptible (*s*), Exposed (*e*) Infected (1), Hospitalized (*h*), Recovered (*r*), Vaccinated (*v*), and Deceased (*d*), and equations given by:

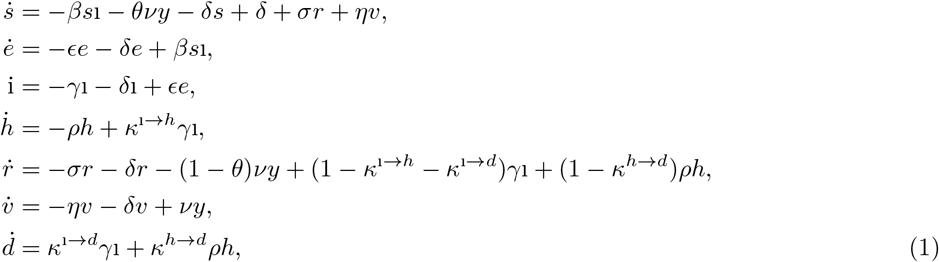

where for a scalar-valued variable *t* ↦ *x*(*t*) we use 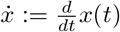 to denote its time-derivative. In the model (1), infected individuals infect susceptible ones with a transmission rate *β >* 0. Individuals become infectious after an incubation period 1*/ϵ*, and they recover at a rate *γ >* 0. After being infected, a fraction of individuals *κ*^1⟶*d*^ ∈ [0, 1] dies and a fraction *κ*^1⟶*h*^ ∈ [0, 1] is hospitalized. The fraction 1 − *κ*^1⟶*d*^ − *κ*^1⟶*h*^ quantifies the individuals who recover without hospitalization. Hospitalized individuals recover at a rate *ρ >* 0. After being hospitalized, a fraction *κ*^*h*⟶*d*^ ∈ [0, 1] of individuals die, while 1 − *κ*^*h*⟶*d*^ recover. Recovered individuals lose immunity at a rate *σ >* 0 thus returning in the susceptible compartment. We denote by *y* the vaccination rate and by *ν* the vaccination efficacy. Individuals are vaccinated regardless of their prior infection history, and we let *θ* ∈ [0, 1] be the fraction of vaccines that is administered to individuals in the *s* compartment, while (1 − *θ*) vaccines are administered to individuals in the *r* compartment. Finally, *δ >* 0 describes the population birth/death rate. The compartmental model corresponding to the dynamical system (1) is illustrated in Figure 7.

**Figure 7:**
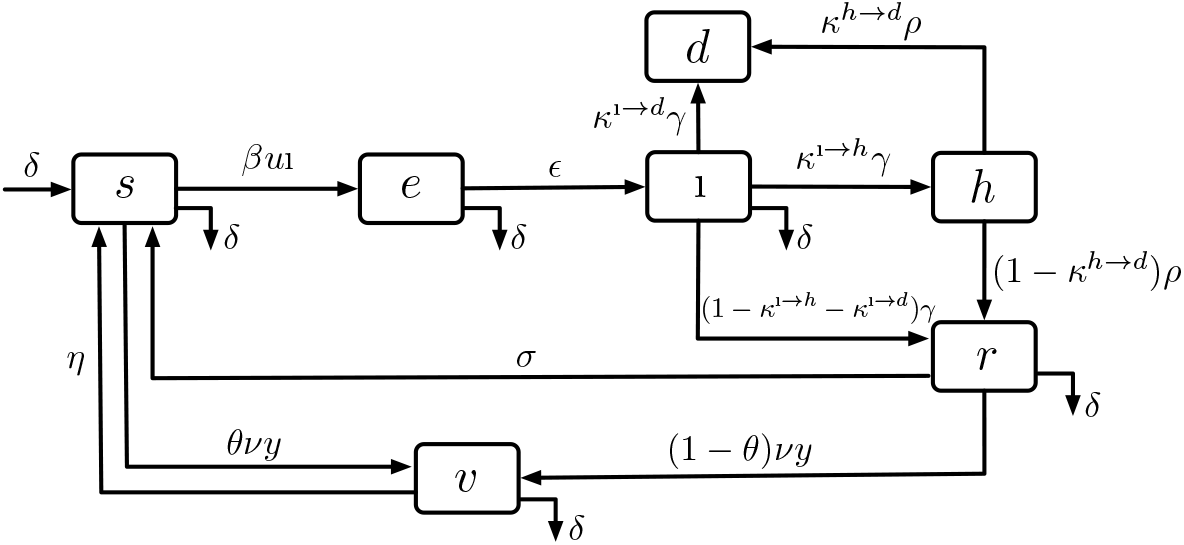
Block diagram of the adopted compartmental model. Model equations are discussed in Methods.

To model NPIs, let *u* be a parameter ranging from 0 to 1 that specifies the level of *permitted* social activity. The special case *u* = 0 corresponds to a full lock-down, while *u* = 1 involves “zero” NPIs (and hence, a return to “normal”). Noticing that different levels of NPIs involve different levels of social interactions (and, consequently, of the transmission rate), the model (1) is updated as follows:

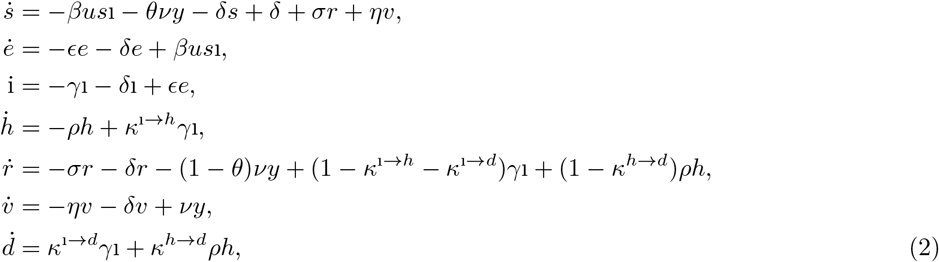

where we note that, whenever *u <* 1, the effective transmission rate is reduced from *β* to *βu* because of the imposed NPIs.

Before proceeding, we emphasize that the parameter *u* ∈ [0, 1] models all NPIs that lead to decreased transmission-relevant contact rates in the population. Currently, it is intractable to disentangle the specific impact that individual interventions have, as multiple complex interventions are introduced simultaneously and the population is reacting in a continuous manner to changing risk perception influenced by divergent policy and messaging at the local, state, and federal level [22].

#### 4.1.2 Optimization-based Controller for NPI

Next, we outline the technical approach that is used for the synthesis of the NPI policy. The prime control objective is to repeatedly update the value of the control variable *u* in order to meet predefined social and health-related metrics and constraints. We begin by presenting a general control formulation that seeks to determine a balance between the permitted level of transmission-relevant contact and the number of infections at the endemic equilibrium. As a particular case, we will then specify the control method to a case where the behavior at the endemic equilibrium is of minor interest and the controller only takes into account the severity of NPIs.

##### Target optimization problem

The proposed technical approach builds upon formulating an optimization problem that captures the desired social and economic metrics, and incorporates constraints related to the largest admissible number of hospitalizations. To this end, let *ϕ* : (0, 1] ⟶ ℝ_≥0_ be a function of the decision variable *u* that models the societal loss induced by the introduction of NPIs [18], including the economic impact of the raised control measures, and/or the societal response to restriction orders. Mathematically, we assume that *u* ↦ *ϕ*(*u*) is a differentiable function with a Lipschitz-continuous derivative [49, 50]. Since societal losses do not increase as NPIs are lifted, we also assume that *u* ↦ *ϕ*(*u*) is a non-increasing function in its domain. Relevant examples of such a functions include:

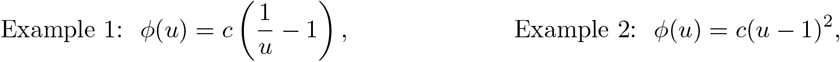

where *c* is a free model parameter; for example, in [18] the choice of *c* is made according to the number of employees that are affected by NPIs in different regions; see Figure 8 for an illustration. For additional remarks on the cost of NPIs we refer the reader to [51, 52].

**Figure 8:**
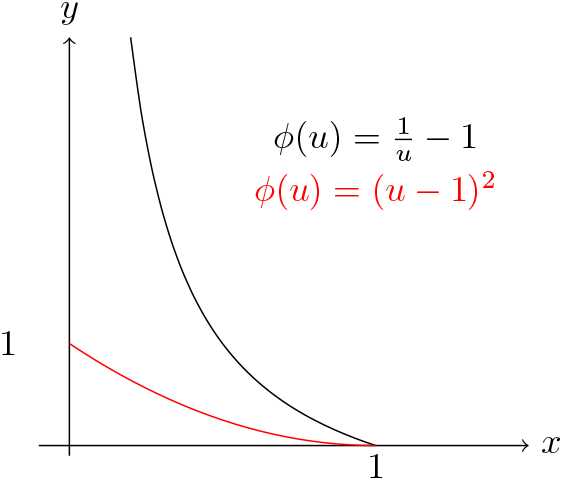
Examples of possible choices of cost functions capturing economic losses and social discomfort induced by different severity levels of the NPI.

To capture the relationship between control variables and the number of infected individuals at the endemic equilibrium (i.e. when *t* = ∞), we denote by *u* ↦ ℱ (*u*) the function that maps the instantaneous values of NPIs *u* to the fraction of infections at the endemic equilibrium of system (2). An exact expression for ℱ can be derived in closed-form, which reads as:

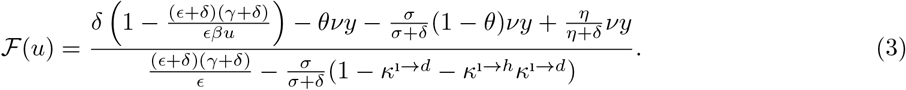

We notice that, when *y* = 0 (i.e. the model does not incorporate vaccinations) and *σ* = 0 (i.e. there is no loss of immunity), the expression (3) simplifies to the well-known input-to-steady state map valid for the SEIR model [53, Section 9.3]:

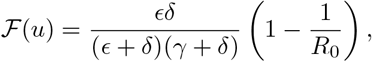

where 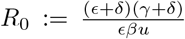 denotes the basic reproduction number. Finally, we let *u ↦ ψ*(ℱ (*u*)) be a loss function that captures the costs associated with the number of infections at the endemic equilibrium.

In this work, we focus on the problem of optimizing the severity of NPIs while maintain the number of COVID-19 hospitalizations under a pre-specified threshold, denoted by *h*_lim_. During the progression of the pandemic, the peak of hospitalizations can be naturally related to the instantaneous level of NPIs as well as to the current state of the model. To this end, we denote by *u* ↦ ℋ (*u*; *x*), with *x* = {*s, e, i, h, r, v*}, the function that maps the instantaneous level of NPIs *u* into the peak of hospitalizations, given the current state of the model *x*. With these definitions in place, we formulate the problem of optimizing the choice of NPIs while guaranteeing that the number of hospitalizations remains below the pre-specified threshold at all times as follows:

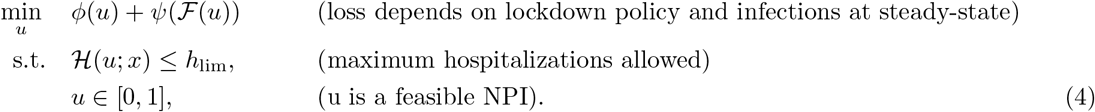

Solutions *u*^*∗*^ of the above optimization problem describe an NPI policy that balances economic and social costs, while ensuring that hospitalizations limits are not exceeded.

##### Controller design for NPIs

To determine optimal NPIs, we propose a feedback controller that leverages information of the current model state, and produces decisions *u* aiming to steer the progression of the epidemic towards a state that coincides with the solution of (4). In words, the controller adapts in real time the NPI so that the objectives set forth in (4) are met. The proposed method relies on feedback-based online optimization techniques for dynamical systems [13, 14, 15, 16], and it is described next.

We begin by defining the set of feasible NPIs, as described in the optimization problem (4), which, for each state *x*, is defined as:

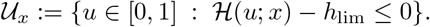

We note that, because the input-to-peak of hospitalization map ℋ (*u*; *x*) is parametrized by the instantaneous state of the system *x*, the set 𝒰_*x*_ is also parametrized by *x*. When the set *U*_*x*_ is (point-wise in time) convex, a suitable control function *t* ↦ *u*(*t*) can be obtained as a solution of the following dynamical system:

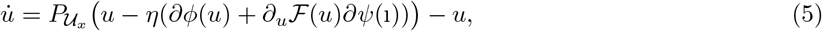

where *η >* 0 is a tunable parameter of the controller, and 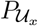 denotes the Euclidean projection operator, namely, given *z* ∈ ℝ^*n*^ and a convex set 𝒰 ⊆ ℝ^*n*^,

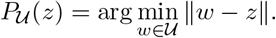

When the set 𝒰_*x*_ is convex and and time-invariant, the controller (5) guarantees two key properties: (i) the trajectories of the dynamical system (2) converge to the set *u* ∈ 𝒰_*x*_; (ii) the feasible set is forward invariant, i.e., if *u*(*t*_0_) ∈ 𝒰_*x*_ for some *t*_0_ ∈ ℝ_≥0_, then *u*(*t*) ∈ 𝒰_*x*_ for all *t ≥ t*_0_ [16, 54]. We note that the controller (5) leverages two types of feedback from the system:

i. it uses the instantaneous fraction of infected individuals i;
ii. it relies on a projection onto the set 𝒰_*x*_, which is parametrized by the instantaneous state of the system.

For these reasons, the control dynamics (5) describe a *dynamic state-feedback controller* for the epidemic model (2). Critically, the controller relies on the knowledge of the maps *u* ↦ ℱ (*u*) and *u* ↦ ℋ (*u*; *x*). While, in general, ℱ (*u*) admits a closed-form expression as in (3), a closed-form expression for ℋ may not be available, since its derivation requires to explicitly solve the dynamics (2) (see [55]). In addition, nonlinearities in ℋ may lead to constraint sets 𝒰_*x*_ that are non-convex [49, 50]. For these reasons, instead of using ℋ we approximate the peak of infections by map 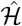, and we modify the controller (5) to take into account projections on approximate sets:

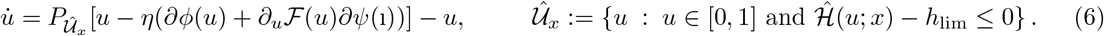

Next, we present a simulation-based technique to estimate the approximate map 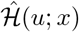.

##### Simulation-based method to estimate ℋ

To estimate the map ℋ, we propose a simulation-based regression method. In particular, we simulate the dynamics (2) with initial condition given by instantaneous model state and for different values of the control parameter {*u*_1_, … *u*_*p*_} (chosen in a neighborhood of *u*(*t*)). Then, we use the output of the simulations to construct the approximate map *u ↦* ℋ (*u*; *x*). Precisely, let {(*ū*_1_, ℋ(*ū*_1_;*x*)), …, (*ū*_*p*_, ℋ(*ū*_*p*_;*x*))} be data-sets obtained by simulating (2) with *x*(0) = *x* with constant inputs {*u*_1_, … *u*_*p*_}, and define:

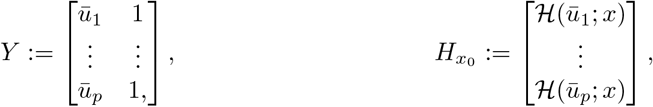

where the subscript *x* is used to highlight the dependence on the model state *x*. Then, we model 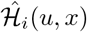 as follows:

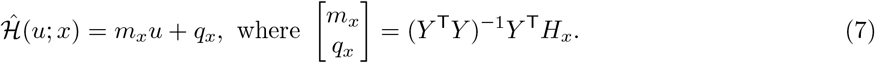

We provide in Figure 10 an illustration of the map ℋ (*u*; *x*) and its approximate counterpart 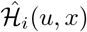, and we conclude by noting that the linear approximation (7) always yields a set 𝒰_*x*_ that is convex.

We also remark that, if ℋ(*u*; *x*_0_) is a non-decreasing function of *u* (see Figure 10 for an illustration) the projection can be computed numerically via line search [49] even for the original set 𝒰_*x*_; however, the simulation-based method explained above yields a procedure that is less computationally burdensome and that works well in practice.

**Figure 9:**
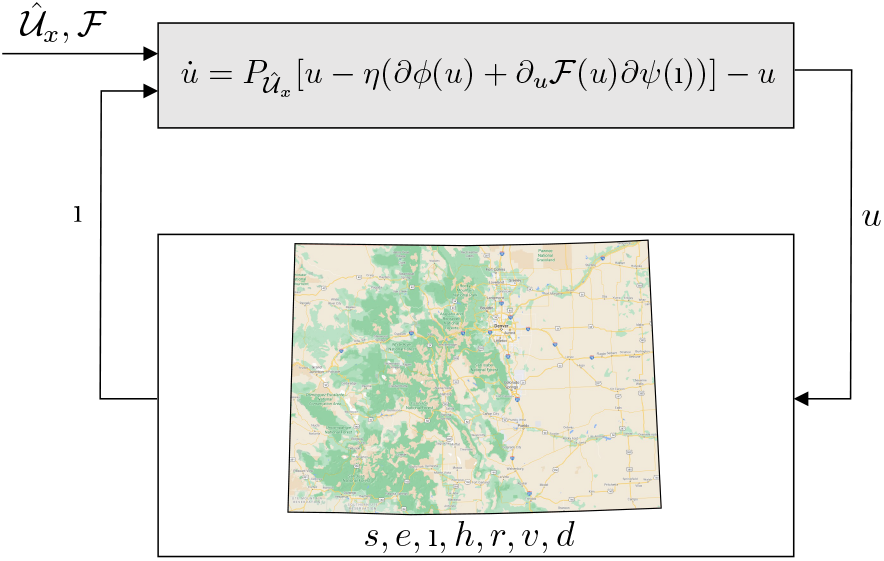
The NPI controller (6) relies on the current fraction of infected people in the region of interest and the set of feasible NPI strategies 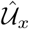; the latter is calculated based on the limit of hospitalizations and a projection of the progression of the infections using the SEIHRVS model. With these inputs, the controller yields a new value for u based on the economic and social metrics defined by the functions ϕ and ψ.

**Figure 10:**
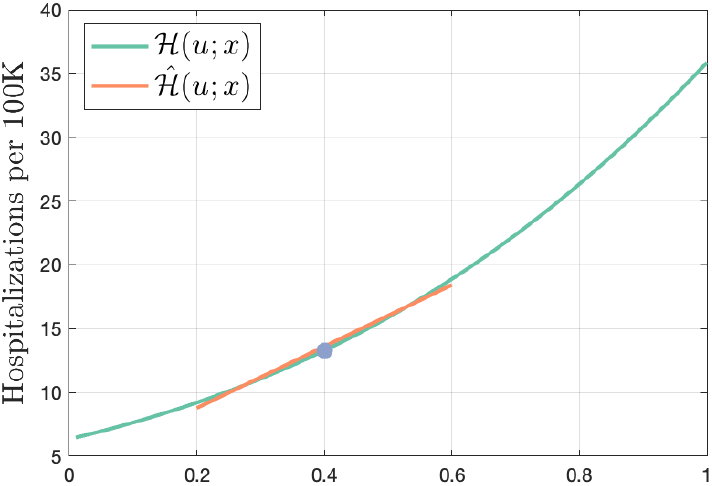
Map ℱ(u, x) estimated via simulations, and approximate map computed according to (7) in a neighborhood of the operating point.

##### Special case: minimizing economic loss

We conclude by discussing a special case in which the number of infections at the endemic equilibrium is disregarded from the loss function. In this case, (4) can be simplified to optimize the severity of the NPIs while maintaining the hospitalizations under a given limit as follows:

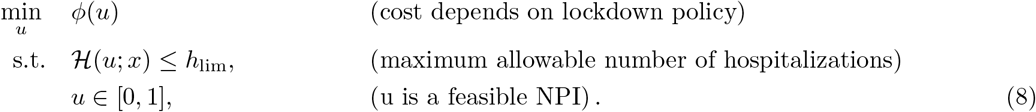

Moreover, we note that, when *ϕ*(*·*) is a monotonically decreasing function in its domain, an optimal NPI policy that solves (8) can be computed in closed form by letting:

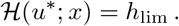

That is, the restrictions on social interactions are lifted to a level where the forecasted amount of hospitalizations will be precisely at the limit *h*_lim_. Although in this case the solution *u*^*∗*^ can be found explicitly, one can still utilize the controller in (5) to adopt a less aggressive approach for lifting the NPIs. In particular, the controller in (5) is simplified in this case as

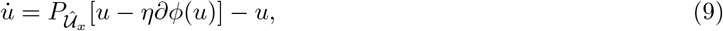

where the set 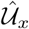 is re-estimated on a daily basis based on the current number of infections, vaccinations, and hospitalizations.

##### Controller stability

We provide a brief remark on the stability of the proposed NPI controller when coupled with the dynamics of the epidemic, as illustrated in Figure 9. We first note that the dynamical system (2) modeling the evolution of the epidemic can be shown to have an endemic equilibrium when the loss of immunity is discarded from the model; the endemic equilibrium can also be shown to be globally asymptotically stable for anu *u* by extending the technical arguments of [53, Section 9.3]. The projected gradient flow (6) can be designed to be globally exponentially stable, as explained in *ψ* [56, 57]; for example, this requires the cost to be strongly convex or to satisfies the Polyak-Lojasiewicz inequality [16].

The stability of the interconnection in Figure 9 can be investigated using arguments from singular perturbation theory [58]; in particular, when the set *U*_*x*_ is constant, one can utilize the procedure explained in [15, 16] to show that there exists a rage of values for the gain *η* that renders the strict optimizers of the optimization problems asymptotically stable [15, 16].

We conclude by noting that the stability of the full dynamical model with the loss of immunity is an open problem; though, we also point out that it is still unclear how to model the loss of immunity of Covid-19 vaccines (the simulations in this document are run for the period of one year, under the assumption that there is no loss of immunity in this time frame). We will investigate this problem in a parallel research effort.

### 4.2 Network model

We consider a model of disease transmission that is organized into a group of geographical subregions, where individuals perform short-term (e.g. daily) inter-region movements. The assumption that movements are short-term refers to scenarios where exposed individuals return to the corresponding region of residence before, eventually, becoming infectious. As an example, Figure 11 illustrates a partition of the state of Colorado into subregions according to LPHA regions.

**Figure 11:**
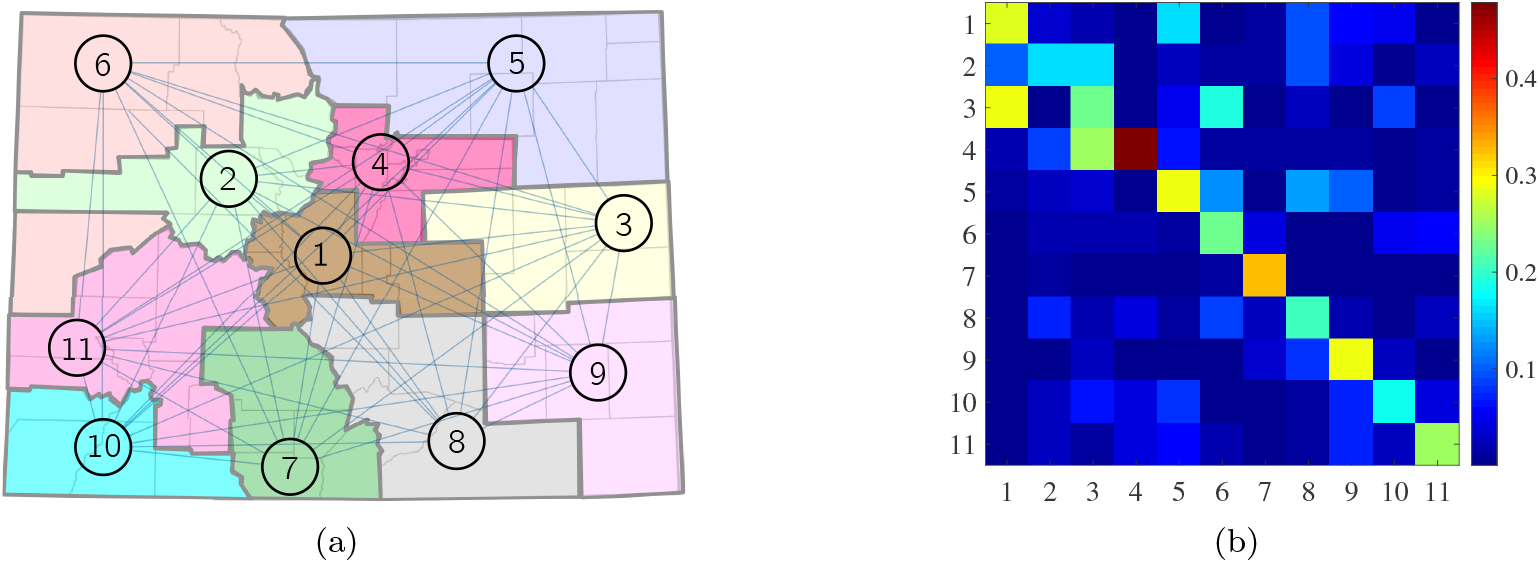
Representative network model of the state of Colorado, USA, according to Local Public Health Agency (LPHA) regions (see also Table 1). (a) Network graph where edges model traveling patterns among the regions. (b) Mobility matrix, where entry (i, j) models the intensity of movements from region j to region i (large values correspond to high interactions between residents of regions). The mobility matrix was estimated by using mobile-phone usage.

The next sections will explain the network SEIHRVS model and the development of the NPI controller.

#### 4.2.1 Network SEIHRVS Model

To model network interactions, we describe the interconnection structure by means of a graph 𝒢 = (𝒱, *ε*) where 𝒱 = {1, …, *N*} denotes the set of nodes (subregions), and *ε* ⊆ 𝒱 *×* 𝒱 denotes the set of edges (interconnections between regions). We model the edge weights by assuming that a fraction *a*_*ij*_ ∈ [0, 1] of residents of region *j* travel to region *i* and interact with its residents. To model the nodes of the graph, we assume that each individual that is a resident of subregion *i* is categorized in one of the seven compartments *s*_*i*_, *e*_*i*_, i_*i*_, *h*_*i*_, *r*_*i*_, *v*_*i*_, *d*_*i*_. Similarly to (2), each state represents the fraction of individuals in the corresponding compartment, with *s*_*i*_ + *e*_*i*_ + i_*i*_ + *r*_*i*_ + *h*_*i*_ + *v*_*i*_ + *d*_*i*_ = 1 at all times. The network model we adopt is as follows:

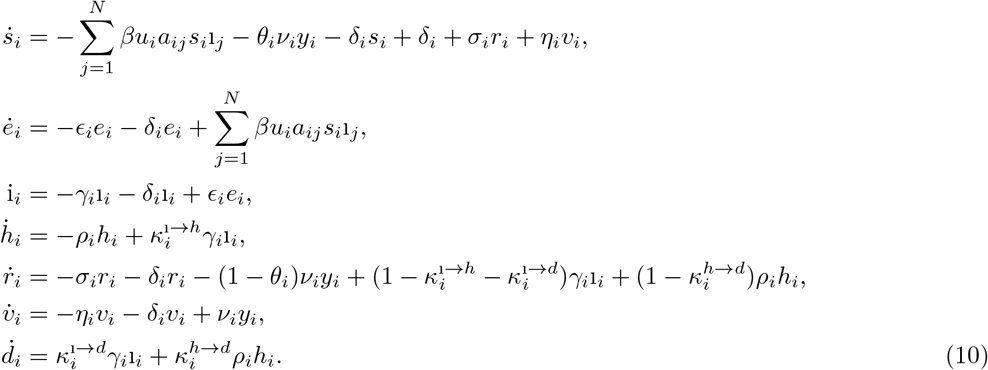

In contrast with (2), the network model (10) incorporates the quantities *a*_*ij*_ denoting the fraction of individuals in region *i* interacting with individuals in region *j*, with Σ *j a*_*ij*_ = 1. Moreover, (10) allows for different levels of NPIs across the regions, where variable *u*_*i*_ ∈ [0, 1] describes the permitted level of social interactions within region *i*.

We conclude by noting that there is increasing evidence to assume that daily movements of individuals highly affect the evolution of an epidemic. For example, [59] found that the regional progress of influenza is much more correlated with the movement of people to and from their workplaces, rather than geographic distances. For these reasons, network models are widely used in the literature to take into account spatial propagation effects and heterogeneous population distributions, see e.g. [60, 61, 62, 63, 64], and [18, 53].

#### 4.2.2 Network Feedback-Based Optimizer for NPIs

We begin by recalling that *u*_*i*_ describes the level of permitted social activity within subregion *i*, and we let *u* = {*u*_1_, …, *u*_*N*_} denote the decision vector describing the level of allowed NPIs in all regions. Let *ϕ*_*i*_ : (0, 1] *→* ℝ_*≥*0_ be a function that captures the societal loss associated with NPIs in region *i*. Moreover, let *u ↦* ℱ_*i*_(*u*) be the function mapping the instantaneous value of NPIs into the fraction of infected individuals in region *i* at the endemic equilibrium. Notice that, in general, ℱ_*i*_(*u*) is a function of the entire vector *u*, as described by the model (10). Moreover, let *u ↦ ψ*_*i*_(ℱ_*i*_(*u*)) denote a loss function that captures the healthcare costs or risks related to the number of infections in the *i*-th region at final time, given the NPIs *u*.

The extension of problem (4) to a multi-area setting can thus be formulated as follows:

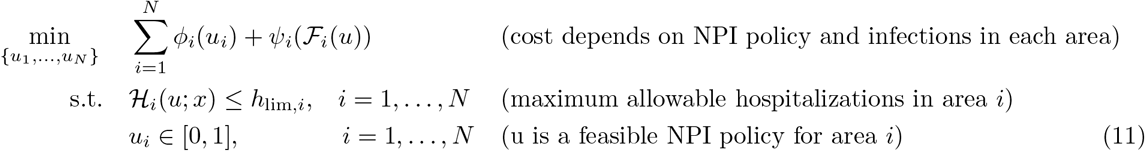

where *h*_lim,*i*_ describes the maximum number of allowed hospitalizations in region *i*, and the function ℋ_*i*_(*u*; *x*) maps the instantaneous level of NPIs *u* into the peak of hospitalizations in the region *i*. Problem (11) formalizes the objective of minimizing a given loss function while guaranteeing that the number of hospitalizations remains bounded by a pre-specified threshold in all regions.

The synthesis of regional controllers follows a similar procedure as presented in Section 4.1.2 for the single-population model. Precisely, define:

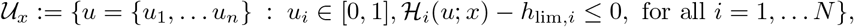

then, the multi-region feedback optimizer reads as:

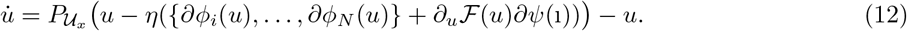

Finally, in order to overcome the lack of knowledge of the maps ℋ_*i*_(*u, x*) and ℱ_*i*_(*u, x*), we use their data-driven approximate counterparts 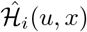 and 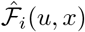 as illustrated in Section 4.1.2.

##### Distributed implementation

Due to the coupling introduced by the dependence of functions ℋ_*i*_ and ℱ*i* on the (entire) vector of control variables *u*, the implementation of a feedback-optimizing controller of the form (12) critically requires a centralized implementation, namely, the step (12) requires full knowledge of variables *u*_1_, …, *u*_*N*_, *h*_lim,1_, …, *h*_lim,*N*_, as well as of the (gradients of) the cost functions *ϕ*_1_, … *ϕ*_*N*_, *ψ*_1_, …, *ψ*_*N*_, and updates the entire controller variables 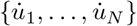. In many cases, such implementation is undesirable because: (i) regions often update NPI policies in an asynchronous fashion, namely, NPI policies may be updated at different time instants at different subregions, and (ii) it is desirable to allow each subregion to design NPIs that are independent of the decisions of the other subregions. For these reasons, we next consider a reformulation of (11) that allows for a distributed implementation. To this aim, we consider an approximation of the functions ℋ_*i*_ that accounts only for the effects of the local NPI policies in area *i*, namely, we approximate the value of the peak of hospitalizations ℋ_*i*_(*u*; *x*) by 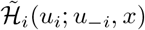, where *u*_*−i*_ = {*u*_1_, …, *u*_*i−*1_, *u*_*i*+1_, …, *u*_*N*_} is treated as a constant parameter. By using this approximation, we redefine the set of feasible NPI policies in subregion *i* as:

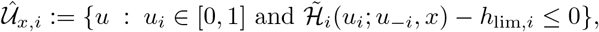

where the map 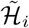 is obtained numerically by using the approach described in Section 4.1.2. By using this reformulation, the proposed distributed controller reads as:

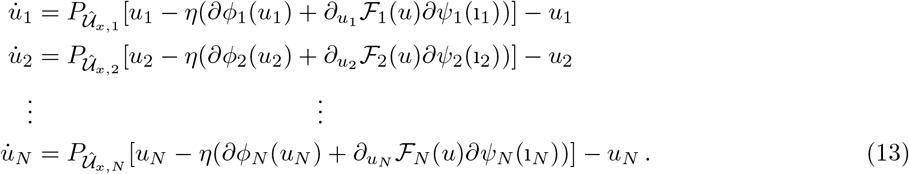

We note that the controllers (13) can be implemented in a distributed fashion. In particular, each region *i* can update its NPI policy *u*_*i*_ by only relying on the knowledge of: (i) the current fraction of infected individual i_*i*_ in region *i*, (ii) the current value of NPI in the network *u*_*−i*_, (iii) an estimate of the partial derivative 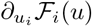, which can be estimated locally at each region by simulating the the model (10), and (iv) the feasible set 𝒰_*x,i*_. A representation of the implementation of the controllers (13) is provided in Figure 12; the figure illustrates the case where the state of Colorado, USA, is partitioned into LPHA regions, and distributed controllers of the form (13) are implemented at each LPHA region. We conclude by noting that, although the controllers are implemented locally in each region, coordination between regions naturally emerges because of the connectivity in the SEIHRVS model.

**Figure 12:**
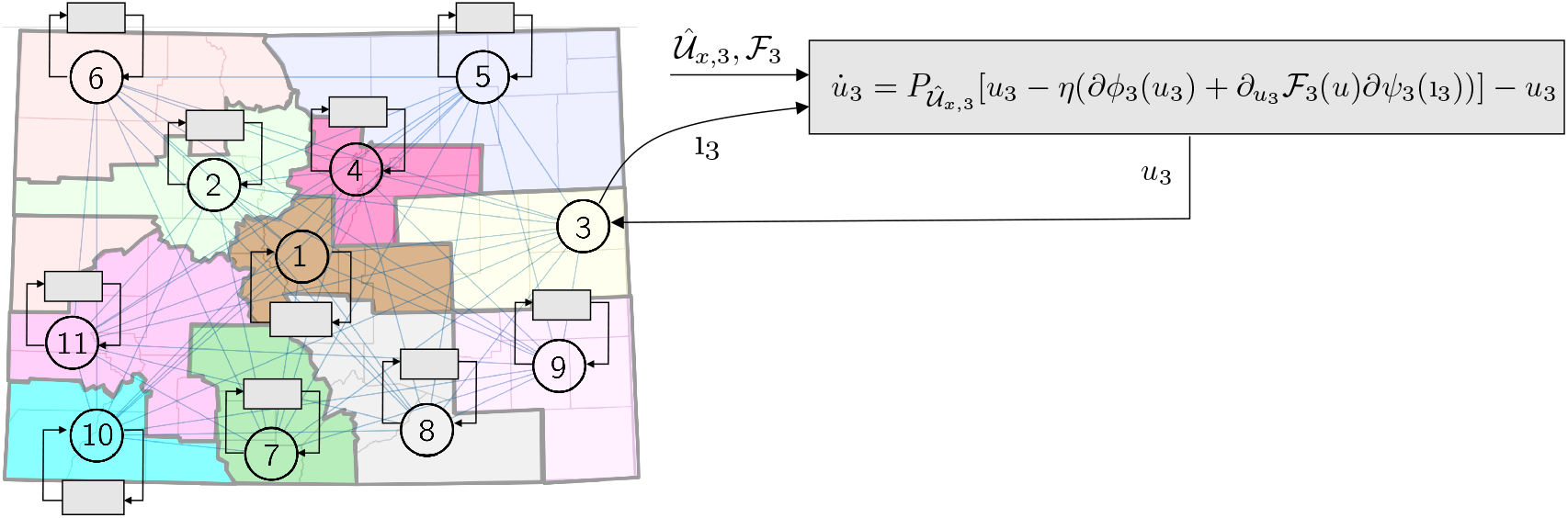
Implementation of the controllers on a per-region basis. The example refers to the state of Colorado, where each region represents a Local Public Health Agency.

###### Remark 1

*We observe that the limit on the hospitalizations h*_lim,*i*_ *in each area does not need to be same during the evolution of the epidemic. If a patient is moved from one hospital in area j to an hospital located in area i, the limits h*_lim,*j*_ *and h*_lim,*i*_ *can be modified to reflect a different number of available hospital beds for residents of the two areas. For example, h*_lim,*i*_ *is updated as h*_lim,*I*_ → *h*_lim,*i*_ *−* 1 *since one hospital bed is occupied by an individual from a different region. The same procedure can be used when some hospital beds are temporarily occupied by tourists*. □

### 4.3 Model Fitting From Data

We organized the model-fitting phase into two main stages. First, we fitted the SEIHRVS model (1) by combining model parameters from [65] with hospitalization data by using a via a prediction-correction algorithm (see Table 2). Second, we used cell-phone usage from SafeGraph (https://docs.safegraph.com/docs/social-distancing-metrics) to estimate the interaction matrix of the network.

**Table 2:** Adopted model parameters.

| Symbol | Value | Description | Source |
| --- | --- | --- | --- |
| $\beta$ | $0.44 \cdot \lambda$ | transmission rate | fitted |
| $\lambda$ | 1.39 | difference in infectiousness symptomatic/asymptomatic | [65] |
| $\theta$ | 0.77 | probability of vaccinating an individual in compartment $s$ | [65] |
| $\delta$ | 0.02965/365 | daily death/birth rate | [65] |
| $\sigma$ | 1/365 | 1/duration of natural immunity | [65] |
| $\eta$ | 1/730 | 1/duration of vaccine immunity | [65] |
| $\epsilon$ | 1/4.2 | 1/latency period | [65] |
| $\gamma$ | 1/9 | rate of recovery | [65] |
| $\kappa^{i \rightarrow h}$ | 0.0143762 | probability of hospitalization after infection | fitted |
| $\kappa^{i \rightarrow d}$ | 0.00262289 | probability of death after infection | [65] |
| $\kappa^{h \rightarrow d}$ | 0.099204 | probability of death after hospitalization | [65] |
| $\rho$ | 1/7.489 | 1/hospitalization period | [65] |
| $y$ | 15,000–25,000 | vaccination rate | |
| $\nu$ | 0.81 | vaccination efficacy | [65] |
| $N_{\text{pop}}$ | 5840795 | state population size, CO, USA | |
| $s(0)$ | 1/1.47 | fraction of Susceptible on 03/01/21 | [19] |
| $e(0)$ | 1/546 | fraction of Exposed on 03/01/21 | [19] |
| $i(0)$ | 1/216 | fraction of Infected on 03/01/21 | [19] |
| $h(0)$ | 1/15936 | fraction of Hospitalized on 03/01/21 | [19] |
| $r(0)$ | 1/4.2136 | fraction of Recovered on 03/01/21 | [19] |
| $v(0)$ | 1/13.1 | fraction of Vaccinated on 03/01/21 | [19] |
| $u(0)$ | 0.21 | level of lockdown on 03/01/21 | [65] |

The fitted model parameters are reported in Table 2. Precisely, model parameters were selected (via a prediction-correction algorithm) to fit hospitalization data for the period from January 18st, 2021, to February 28th, 2021. State-wide hospitalization data was obtained from EMResources and regional-level data came from Colorado COVID Patient Hospitalization Surveillance [21, 65]. This time-interval was selected due to a steady decline in hospitalizations observed over this period, thus overcoming the need to account for varying transmission rates during model-fitting. Validation was carried out by comparing the evolution of the parameterized model with hospitalization data, and by assessing performance through a mean squared error. For the parametrized model, the mean squared error associated with hospitalizations and infections was less than 10% over the entire dataset. The model was initialized on 1/18/21 by using infection data from [19]. The state of the pandemic on 03/01/21 was computed by initializing the model on 1/18/21 and by simulating the evolution of the epidemic with constant vaccination rate of *y* = 12, 000 vax/day and vaccination efficacy *ν* = 0.9.

Cell-phone data usage was used to estimate the mobility matrix. Data was obtained from SafeGraph, in the form of counts of cell phone accounts that normally reside in county *i* and traveled to county *j* on a specific date, have been averaged over the time interval 03/01/20–12/31/2020.

All computational analyses and the fitting of data were performed using MATLAB and the corresponding optimization toolbox. Our Montecarlo simulations suggest that the outcomes of our simulations are robust to variations of the parameter, confirming their validity to describe reality. Note that the model describing the epidemic spread is highly nonlinear and therefore potentially sensitive to parameter perturbations.

## Data Availability

The raw data supporting the conclusions of this article will be made available by the authors, without undue reservation.

## Acknowledgments

The EMResource data on hospital census is publicly available on the website https://covid19.colorado.gov/data. The dataset used in this study is publicly available at https://docs.safegraph.com/docs/; the authors would like to thank David Jacobson (Innovation Response Volunteers, Boulder, CO) for making the data available for research use.

